# Lynch syndrome in the general population: low clinical utility of genomic screening for *PMS2* pathogenic variants

**DOI:** 10.1101/2025.02.03.25321630

**Authors:** Kelly M. Schiabor Barrett, Catherine Hajek, James Lu, Kate Lynch, Leigha Senter, Amy C. Sturm, Chad R. Haldeman-Englert, Jeremy Cauwels, Douglas Stoller, Christopher N. Chapman, C. Anwar A. Chahal, Daniel P. Judge, Douglas A. Olson, Joseph J. Grzymski, William Lee, Elizabeth T. Cirulli, Alexandre Bolze

## Abstract

**Purpose:** Screening for Lynch syndrome in the general population aims to prevent and detect cancers early. However, current clinical guidelines for those with pathogenic variants are largely based on studies of patients with cancer or strong family history. The objective of this study was to determine the risk of cancer associated with pathogenic variants in *MLH1*, *MSH2*, *MSH6*, or *PMS2* in the general population.

**Methods:** This retrospective case-control study utilizes Helix Research Network^TM^ data from 216,095 participants across nine US health systems. Variant interpretation was performed following the ACMG-AMP guidelines. Clinical diagnoses were identified from electronic health records for 11 cancer types associated with Lynch syndrome, including colorectal and endometrial cancers.

**Results:** Individuals with pathogenic variants in *PMS2* had a small increase in risk for all 11 Lynch syndrome-associated cancers with Hazard Ratio (HR) of 1.6 (95% CI: 0.9-2.7) compared to those without a pathogenic variant. The increase in risk was also small when restricting the analysis to colorectal cancer with HR of 4.4 (2.1-9.3). This increase in colorectal cancer risk was predominantly observed after age 60, 10 years after USPSTF guidelines recommend starting colonoscopies for the average population. Up to age 60, 2.3% of individuals with *PMS2* pathogenic variants were diagnosed with colorectal cancer, closer to the general population (0.5%) than those with *MLH1* (44.4%), *MSH2* (33.7%) or *MSH6* (6.6%) pathogenic variants. For 100,000 individuals screened for *PMS2*, 875 additional colonoscopies would be performed, which would prevent or detect early 2.9 colorectal cancers.

**Conclusion:** This analysis across nine health systems highlights the low clinical utility of genomic screening for *PMS2* variants in the general population.

## Introduction

Lynch Syndrome (LS) is due to pathogenic variants in mismatch repair (MMR) genes: *MLH1*, *MSH2*, *MSH6* and *PMS2 ^1^*. LS is the most common colorectal cancer syndrome and is screened for as part of population health programs because of the high penetrance as well as the existence of guidelines for intervention and prevention ^2, 3^. For example, surveillance strategies recommended by the National Comprehensive Cancer Network® (NCCN®) for those with a *MLH1* or *MSH2* pathogenic variant are to start high-quality colonoscopy at age 20-25y and repeat every 1-2y ^4, 5^, and to consider using daily aspirin^4^. For individuals with a *MSH6* or *PMS2* pathogenic variant, NCCN® recommends starting high-quality colonoscopy at age 30-35y, repeating every 1-3y ^4^ (**Supplementary Table 1**). Comparatively, the US Preventive Services Task Force recommends starting colorectal cancer screening at age 45-50y for all individuals, repeating colonoscopy every 10y^6^.

The gene specific NCCN® recommendations are largely based on risk estimates from studies of patients who were sequenced because of a personal or family history of colorectal cancer, such as the Prospective Lynch Syndrome Database (PLSD) ^7–9^. In its latest data release, the PLSD study included 8,500 participants who all had pathogenic variants in *MLH1* (36.8% of the 8,500 participants), *MSH2* (37.3%), *MSH6* (19.4%), or *PMS2* (6.5%). However, the study did not contain a control population, in terms of either outcomes in relation to standard of care prevention or of individuals without pathogenic variants in MMR genes. Disease risk estimates derived from cohorts of patients already diagnosed with cancer or with established high risk for cancer may not be adequate when returning results to an unselected individual in a population genomic screening context.

A remaining key question for healthcare providers, including primary care physicians, gastroenterologists, and oncologists, is whether the current surveillance strategy recommendations for Lynch syndrome are relevant in the context of population-based genomic screening. The aims of our study are: (i) to identify the prevalence of individuals with pathogenic variants in the MMR genes in the general population, (ii) to measure the rate of cancer diagnoses and determine the risk associated with pathogenic variants in these genes in the general population, and (iii) to intersect this data with current surveillance recommendations.

We will study these aims across the Helix Research Network, with data from 216,095 participants from 9 health systems in the United States. Retrospective analyses of all-comer clinicogenomic cohorts, where all participants are sequenced without qualifying criteria, provide an opportunity to estimate and compare population-based risks for individuals both with and without pathogenic variants, in the context of the standard of care.

## Methods

### Study design and participants

This is a retrospective and observational clinico-genomic analysis of a prospectively designed study. All participants were adults enrolled in the Helix Research Network^TM^ (HRN) study, which is a protocol open to general patient populations at various U.S. healthcare organizations. All participant data analyzed for this publication came from nine U.S. Health Systems participating in the HRN study. The studies included under the HRN protocol are: Ohio State Genomic Health (The Ohio State University Wexner Medical Center), GeneConnect (Cone Health), ImagineYou (Sanford Health), DNA Answers (St. Luke’s University Health Network), the Genetic Insights Project (Nebraska Medicine), the Healthy Nevada Project (Renown Health), In Our DNA SC (Medical University of South Carolina), myGenetics (HealthPartners), and the Gene Health Project (WellSpan Health). Study protocols were reviewed and approved by their respective Institutional Review Boards (projects 956068-12 and 21143). All participants provided informed consent via a digital consent process prior to participation. Direct identifiers were removed from the research dataset to protect participant privacy. Participation in these studies was at no cost to participants. For this analysis, data from 216,095 participants with linked Electronic Health Records (EHR) and Exome+® sequencing data were included.

### Comparison with SEER database

Analysis used data extracted from SEER explorer (https://seer.cancer.gov/statistics-network/explorer/application.html) with the following criteria on 18 April 2025 ^10^:

- Cancer site: colon and rectum (including appendix).
- Statistics to explore: Prevalence. Complete.
- Compare by age at prevalence.
- Precision: 0.001.

### Genetics and Variant interpretation

#### Genetic data

Saliva or blood samples were collected from participants and underwent Exome+® sequencing at Helix between February 2018 and February 2025. The Exome+® assay includes a clinical exome, which is used for return of clinical results for CDC Tier 1 genes, including Lynch syndrome genes, as previously described^2^. Variants in exons 11-15 of *PMS2* are more difficult to call due to the existence of a pseudogene. Helix developed a tailored clinically-validated bioinformatics pipeline to identify variants in these exons, which are then confirmed by an orthogonal assay. To maximize consistency and reproducibility, we opted to exclude exons 11-15 of *PMS2* from our analysis for the following reasons: (i) some older samples were not analyzed with this pipeline, and (ii) the larger population-based studies, which we were aware of, looking at *PMS2* (e.g. UK Biobank and All of Us) did not include variants in these exons. At the time of our analysis, only 1 pathogenic large deletion in *EPCAM* going into the promoter of *MSH2* was identified and confirmed. This large deletion affected 1 participant, and we decided to not include it in the analysis given it was a N of 1. Genotype processing for Helix data was performed in Hail 0.2.115-10932c754edb.

#### Variant Interpretation

Variant interpretation for the four mismatch repair (MMR) Lynch syndrome genes (*MLH1*, *MSH2*, *MSH6*, and *PMS2*) was completed for the entire HRN cohort (N=216,095) using a two-step approach. First, a variant was considered pathogenic if it carried a known and well-established clinical pathogenic interpretation (i.e., no VUS or benign interpretations present in ClinVar across high volume laboratories, using search strings [’ClinGen’, ‘Quest’, ‘Sema4’, ‘Natera’, ‘Invitae’, ‘All of Us’, ‘Baylor’, ‘GeneDx’, ‘Ambry’, ‘LapCorp’, ‘Color’, ‘Myriad’, ‘Brigham’] and/or a likely pathogenic or pathogenic interpretation by the InSiGHT Hereditary Colorectal Cancer/Polyposis Expert Panel (InSiGHT VCEP). For all remaining variants, ACMG-AMP variant interpretations were completed programmatically following the gene-specific scoring recommendations from the InSiGHT VCEP. Data from individual case studies as well as patient-specific information such as presenting symptoms or family history were not considered for these interpretations. All variants seen in HRN, relevant annotations, scoring by data category, and resulting interpretation based off point totals (pathogenic[>5], higher scoring VUS[3-5], lower scoring VUS[-1 to 2], benign[< -1]) are available in **Supplementary Table 2.** Variant annotations were made based on the MANE transcript for each gene *(MLH1*: NM_000249.4, *MSH2*: NM_000251.3, *MSH6*: NM_000179.3, *PMS2*: NM_000535.7) and leveraged the following tools: VEP-104^11^, GnomADv3^12^, REVEL^13^, SpliceAI^14^, ClinVar database (accessed: 03/10/2025), and for *MSH2* functional scores from MAVE (urn:mavedb:00000050-a)^15^, and case-control (PS4) data was obtained from systematic variant-level association tests internally-calculated using clinicogenomic data from UK Biobank and All of Us cohorts (phenotypes leveraged from phecodeX map include: CA_101.41, CA_106.21 for colorectal and endometrial cancers, respectively^16^). The number of HRN participants harboring pathogenic variants in each gene is presented in **Supplementary Table 3**.

### Phenotypes

Electronic health records data were available for all participants included in the study. EHR data were transformed into the Observational Medical Outcomes Partnership (OMOP) Common Data Model (CDM) version 5.4. EHR data accessible included both current and prior history of disease, encompassing a mean of 12.1 years (median: 11.1 years, IQR: 11.3 years) of EHRs per patient. A total of 11 different types of cancer were analyzed for this study, based on prior literature on Lynch syndrome including a comprehensive report from the prospective Lynch syndrome database (see their Table 1 for the list of cancer types) ^9^. The 11 cancer types included were: colorectal, endometrial and uterine, ovarian, kidney and ureter, small bowel, bladder, stomach, pancreas, biliary tract, brain, and prostate cancers. All the OMOP condition concept ids used to extract data from the EHR are in **Supplementary Table 4**.

**Table 1:** Demographics from the Helix Research Network data.

| Demographics table | Number of participants (% of total) |
| --- | --- |
| Participants with genetic and phenotype data available | 216,095 |
| Sex | F: 154,900 (71.7%)<br>M: 61,195 (28.3%) |
| Mean age in 2024, y (min - max) | 52.3 (18 - 106) |
| Genetic similarity | Africa: 10,212 (4.7%)<br>Americas: 13,998 (6.5%)<br>East Asia: 3,707 (1.7%)<br>Europe: 181,105 (83.8%)<br>South Asia: 1,620 (0.7%)<br>Other: 5,453 (2.5%) |
| with a colorectal cancer dx | 1,313 (0.61%) |
| with a endometrial/uterine cancer dx | 783 (0.51%*) |
| with an ovarian cancer dx | 476 (0.31%*) |
| with a kidney/ureter cancer dx | 698 (0.32%) |
| with a bladder cancer dx | 590 (0.27%) |
| with a stomach cancer dx | 98 (0.05%) |
| with a pancreas cancer dx | 261 (0.12%) |
| with a small bowel cancer dx | 53 (0.02%) |
| with a biliary tract cancer dx | 88 (0.04%) |
| with a brain cancer dx | 339 (0.16%) |
| with a prostate cancer dx | 2,846 (4.7%**) |
| avg EHR lookback, y | 12.1 |

\* percentage based on overall number of female participants
\*\* percentage based on overall number of male participants

### Clinical guidelines

We used the NCCN® guidelines^4^ as a basis to assess (i) whether current recommendations were appropriate given the risk of cancer observed, and (ii) count the number of colonoscopies that would be added or subtracted if guidelines were to be changed. We focused on colorectal and endometrial cancers, the two main cancers associated with Lynch syndrome. The recommendations for surveillance and prevention strategies for colorectal cancer are summarized in **Supplementary Table 1**. For endometrial cancer, the recommendations emphasize education surrounding symptoms and speak to possible prevention opportunities such as hysterectomies, biopsies, and ultrasounds that can be considered.

### Survival analysis, hazard ratios, and statistical tests

Kaplan Meier survival curves were done using the KaplanMeierFitter function from the Lifelines python library. The lifelines package was used for time to event analyses including cumulative incidence plots, log rank test, and cox proportional hazard calculations^17^. For time to event analyses, the earliest age at relevant diagnosis or current age (in 2024) was determined for each participant.

### Genomic screening evaluation model

We defined preventable adverse outcomes as a diagnosis of colorectal cancer (CRC) before age 60.

N_adverse_outcomes_prevented_MLH1 = N_with_P_in_MLH1_per100000 x CRC_risk_by_age_60

Number Needed to Screen (NNS) was defined as the number of participants who must be enrolled in a population genomic screening program to prevent 1 adverse outcome.

NNS_MLH1 = 1/(fraction_with_P_in_MLH1 x CRC_risk_by_age_60) = 1/0.000125 = 8,000

NNS_MSH2= 1/(50/216,095 x 0.337)= 1/0.0000780= 12,820

NNS_MSH6= 1/(325/216,095 x 0.066)=1/0.0000993 = 10,070

NNS_PMS2 = 1/(270/216,095 x 0.023) = 1/0.0000287= 34,840

NNS_Lynch_4genes = 1/(0.000125 + 0.0000780 + 0.0000993 + 0.0000287) = 3,020

## Results

### Rates of colorectal cancer in HRN match expectations based on SEER data

We studied Helix Research Network™ data from 216,095 participants across nine health systems. For each participant, genetic data from Exome+® sequencing as well as phenotype data from electronic health records were available (on average, EHR data had a 12.1 years lookback). The average age in 2024 of participants was 52.3 years old, and 37.2% (n=80,366) were 60 or older at the time of analysis (**Table 1**). There were no inclusion or exclusion criteria to participate in the study based on the presence or absence of a personal or family history of cancer. We therefore hypothesized that the prevalence of colorectal cancer in our participant population would be similar to the prevalence of colorectal cancer in the general US population. To test our hypothesis, we calculated the expected prevalence of colorectal cancer based on the prevalence of colorectal cancer reported in the SEER explorer database, using the current age and sex demographics of HRN. Through this exercise, we expected 1,321 colorectal cancer cases. In HRN, we identified 1,313 participants (0.61% of the cohort) with at least one diagnosis of colorectal cancer in their EHR (**Supplementary Table 5**), closely matching the expectations from SEER and showing no clear bias related to colorectal cancer diagnosis compared to the average population in the US.

### *PMS2* pathogenic variants account for 40% of individuals with Lynch Syndrome in the general population

Variant interpretation for MMR LS genes – *MLH1*, *MSH2*, *MSH6* and *PMS2* – was done using well-established clinical interpretations and according to ACMG-AMP criteria and following recommendations from InSiGHT Hereditary Colorectal Cancer/Polyposis Expert Panel (see **Methods**). The list of all variants identified in these genes, the annotations, evaluations of each criterion and final score and pathogenicity assignments are provided in **Supplementary Table 2**. We found that 706 participants (0.33% or 1 in 310) had one pathogenic (P/LP) variant in one of the 4 genes. The majority had either a pathogenic variant in *MSH6* (n=325) or *PMS2* (n=270), while fewer had a pathogenic variant in *MLH1* (n=61) or *MSH2* (n=50) (**Supplementary Table 3**). These gene-based proportions are opposite of the composition of the PLSD, where the majority of individuals were found to harbor pathogenic variants in *MLH1* and *MSH2*. These differences suggest that understanding LS-related risk for *MSH6* and *PMS2* in a screening context is of great importance.

### Risk of diagnosis of a Lynch syndrome related cancer is gene-dependent and lowest for *PMS2* pathogenic variants

To investigate the impact of pathogenic variants in the different MMR genes, we extracted the date of first diagnosis recorded for any of 11 cancers known to be relevant for Lynch syndrome (**Methods**, **Supplementary Table 4**) ^9^. The prevalence of 8 of these 11 cancers is very low (<0.5%) in our unselected population. Therefore, we decided to do a time-to-first diagnosis analysis for colorectal cancer alone, for endometrial cancer alone, and one where we combined all known 11 relevant cancers together (**Figure 1, Supplementary Table 6**)^9^. As expected, those with a pathogenic variant in *MLH1* or *MSH2* were at much higher risk to develop any of the 11 cancers compared to those without a pathogenic variant or VUS in the four genes: Hazard Ratio_MLH1_11cancers_ = 14.1 (95% Confidence Interval: 8.5-23.5) and HR_MSH2_11cancers_ = 15.4 (95%CI: 9.6-24.9). The increased risk was even stronger when looking more specifically at colorectal cancer, HR_MLH1_colo_ = 57.7 (32.6-101.9) and HR_MSH2_colo_ = 62.2 (36.0-107.5). Those with a pathogenic variant in *MSH6* had an intermediate risk for any of the 11 Lynch syndrome relevant cancers HR_MSH6_11cancers_ = 4.2 (3.1-5.7). The strongest risk related to *MSH6* pathogenic variants was for endometrial/uterine cancers in female participants HR_MSH6_endo_ = 13.8 (8.3-23.0) (**Figure 1**).

**Figure 1:**
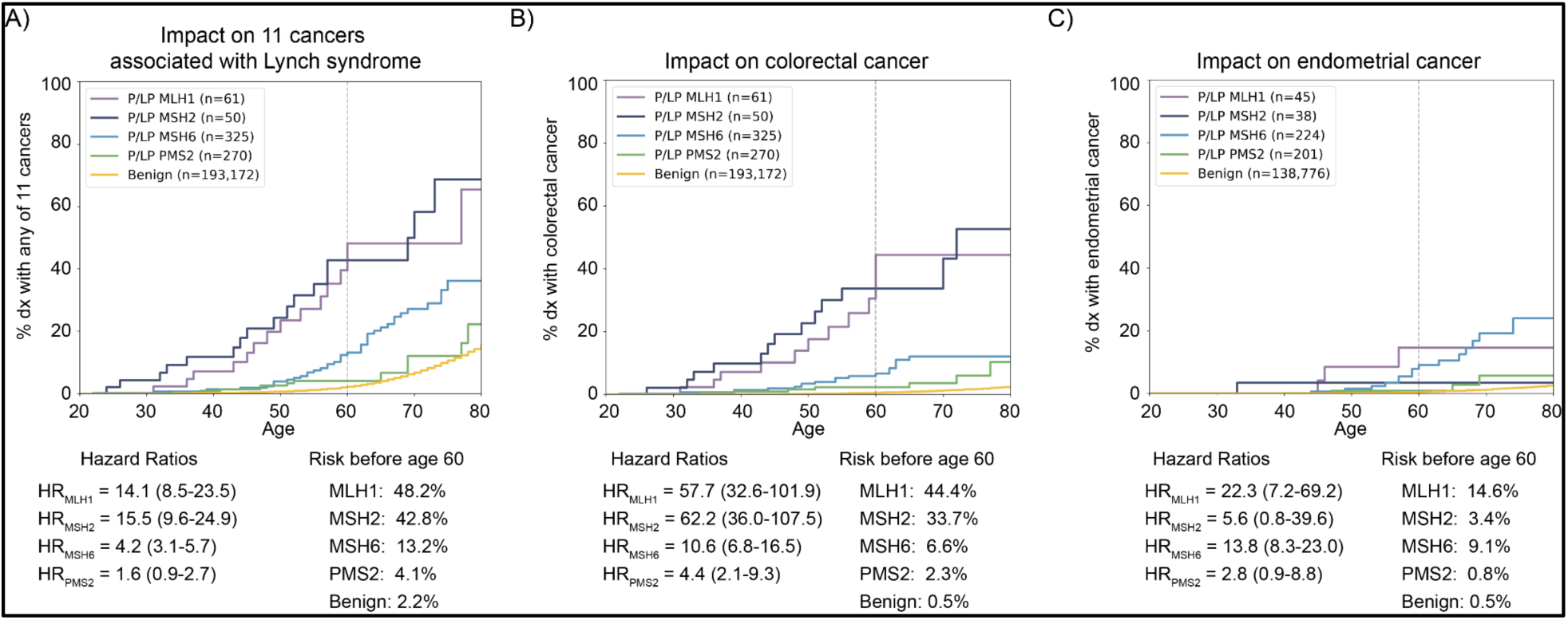
Gene by gene cumulative incidence of age at first Lynch syndrome-related cancer diagnosis across individuals harboring pathogenic variants from the Helix Research Network. Gene-level results for those harboring pathogenic (P/LP) variants (*MLH1* in purple, *MSH2* in blue, *MSH6* in light blue and *PMS2* in green) against those with a benign or no variant (yellow) are shown for: A) All 11 Lynch syndrome cancers: colorectal, endometrial and uterine, ovarian, kidney and ureter, small bowel, bladder, stomach, pancreas, biliary tract, brain, and prostate. B) Colorectal cancer. C) Endometrial cancers in female participants. Hazard ratios (with confidence intervals) as well as the percentage of individuals in each genetic group with a relevant diagnosis at or before age 60 are shown below each panel.

On the other hand, pathogenic variants in *PMS2* were not significantly associated with an increased risk of developing any of the 11 LS-relevant cancers compared to those without a pathogenic variant or VUS variant in an MMR gene (P=0.08, log-rank test). *PMS2* was only associated with colorectal cancer; however, the increased risk and onset of this risk were smaller and later compared to the cancer risks observed for the other MMR genes (HR_PMS2_colo_ = 4.4 (2.1-9.3), **Figure 1**). By age 70, the penetrance of *PMS2* pathogenic variants was 3.6% for colorectal cancer and 12.1% for all 11 LS-associated cancers combined. These results showed that frequency of pathogenic variants in the general population and risk of cancer were inversely correlated.

### Clinical implications of heterogenous risk profiles across Lynch syndrome genes

Our third aim was to measure the implications of returning genomic screening results for Lynch syndrome in a population-based context. The US Preventive Services Task Force (USPSTF) recommends colorectal cancer screening for adults 50-75 yo (grade A recommendation, substantial net benefit), and for those 45-49 yo (grade B recommendation, moderate benefit)^6^. The recommended frequency of screening varies based on the type of test; it is every 10 years for a colonoscopy. Given that the goal of screening is to identify cancers early and that the official grade A recommendation is to perform a colonoscopy every 10 years starting at age 50, we focused our prevention analysis on colorectal cancer diagnoses up to age 60 (as cancers diagnosed after would or could have been identified following a routine screening regimen). By age 60, the risk of developing colorectal cancer was 2.3% for those with a *PMS2* pathogenic variant, modestly higher than the average population (0.5%), yet less than half the risk seen in those with *MSH6* (6.6%) and substantially below rates for both *MLH1* (44.4%) and *MSH2* (33.7%) (**Figure 1**). By age 60, the risk of developing any of the 11 cancers associated with Lynch syndrome was most similar for those with a *PMS2* pathogenic variant (4.1%) compared to the average population (2.2%), whereas this risk is much higher for those with a *MLH1* pathogenic variant (48.2%), *MSH2* pathogenic variant (42.8%) or *MSH6* pathogenic variant (13.2%) (**Figure 1**).

To assess the benefits of screening for the different Lynch syndrome genes in the general population, we calculated the number needed to screen (NNS), which is the number of participants who must be enrolled in a population genomic screening program to prevent one adverse outcome, which was defined as colorectal cancer diagnosed before the age of 60 (**Methods**). The NNS varies substantially by gene, ranging from 8,000 for *MLH1* up to 34,840 for *PMS2*, but it is lowest when modelling a program that identifies LS across all four MMR genes (see **Table 2** for all calculations). On the other hand, testing for *PMS2* in the general population would lead to a significant increase in the number of colonoscopies per 100,000 patients for a low rate of adverse outcomes prevented when following recommendations from current NCCN guidelines. Testing for *PMS2* would lead to 875 additional colonoscopies to be performed per 100,000 patients and would prevent 2.9 adverse outcomes, while testing for *MLH1* would lead to 462 additional colonoscopies and would prevent 12.4 adverse outcomes (**Table 2**). In other words, this model showed that it was eight times more efficient to screen for *MLH1* than *PMS2* in the general population.

**Table 2:** Number of potential adverse outcomes prevented and additional colonoscopies per gene tested in the general population.

| Gene | N of individuals with P/LP vars per 100,000 people | CRC risk by age 60 | N of preventable adverse outcomes identified before age 60 per 100,000 people | NNS | Age start colonoscopies in years* | Freq of colonoscopies* | N colonoscopies per individual by age 60 | Additional colonoscopies per individual with a P/LP var | Additional colonoscopies per 100,000 people screened | Ratio adverse events prevented per additional colonoscopy |
| --- | --- | --- | --- | --- | --- | --- | --- | --- | --- | --- |
| No P/LP | 99,673 | 0.005 | NA | NA | 50 | every 10y | 1 | Reference | Reference | NA |
| *MLH1* | 28 | 0.444 | 12.4 | 8,000 | 25 | every 2y | 17.5 | 16.5 | 462 | 0.027<br>(1 per 37) |
| *MSH2* | 23 | 0.337 | 7.8 | 12,820 | 25 | every 2y | 17.5 | 16.5 | 380 | 0.021<br>(1 per 49) |
| *MSH6* | 150 | 0.066 | 9.9 | 10,070 | 35 | every 3y | 8 | 7 | 1,050 | 0.0094<br>(1 per 110) |
| *PMS2* | 125 | 0.023 | 2.9 | 34,840 | 35 | every 3y | 8 | 7 | 875 | 0.0033<br>(1 per 300) |
| *MLH1* + *MSH2* + *MSH6* + *PMS2* | 326 | NA | 33 | 3,020 | NA | NA | NA | NA | 2,767 | 0.012<br>(1 per 84) |

\* we used the latest starting age, and the least frequent repeat of colonoscopies within the windows recommended by the NCCN guidelines in order to have conservative estimates.

## Discussion

No clinical guidelines exist today for population genomic screening programs including which genes should be included. Our results show attenuated clinical impact for pathogenic variants in *PMS2* at the population level, and can be utilized toward the development of evidence-based guidelines. Our study controlled for potential biases that could have led to artificially low penetrance. First, we found that the rate of colorectal cancer diagnoses observed in our cohort were close to what was predicted using SEER data. Second, we restricted our analysis to exons 1 to 10 of *PMS2* to avoid false positives caused by a pseudogene with high sequence similarity to exons 11-15 of *PMS2.* Third, the high penetrance and increased clinical risk associated with pathogenic variants in *MLH1*, *MSH2*, and even *MSH6* in our cohort suggest that the low penetrance of *PMS2* pathogenic variants is not the result of studying a ‘healthier population’ compared to the general population. Moreover, our results were consistent with two recent studies that assessed the impact of Lynch syndrome in a population-based setting. In the UK Biobank, Table S2 of Fummey et al. showed that 1.3% of individuals between 60-64 yo with a pathogenic variant in *PMS2* had been diagnosed with colorectal cancer compared to 1.0% for non-carriers^18^. In All of Us, Park et al. showed in Figure 1B that the rate of colorectal cancer diagnosis was the same between those with a *PMS2* pathogenic variant and non-carriers until age 60 ^19^.

In the context of clinical care, our analysis focused on colorectal cancer because the NCCN® has strong recommendations for colorectal cancer prevention in those with a pathogenic variant in a MMR gene. We centered our analysis on events and colonoscopies before the age of 60, which was 10 years after the USPSTF guidelines recommend starting colonoscopies for the general population^6^. Our analysis showed that screening 100,000 individuals in the general population for *PMS2* could prevent 2.9 adverse outcomes (early diagnosis of colorectal cancer) through 875 additional colonoscopies. Taken together, population genomic screening programs for LS that include all four genes have the potential to prevent more cancer events (e.g. via NNS statistics). However, the heterogeneity in the ratio of preventable adverse events per colonoscopies by gene highlights the core tension of expanding versus curating population screening programs in terms of what genes are being included in return of results. For individuals, the question also arises whether it is worth the efforts (financial cost, pain, risk of a procedure etc.) to start screening 15 years earlier and do 7 or more additional colonoscopies when the risk of colorectal cancer is only 2.3% by age 60 (**Figure 1**, **Table 2**). One solution to reconcile these tensions may be to amend the existing NCCN guidelines to provide gene-specific recommendations when pathogenic variants in MMR genes, especially in *PMS2*, are identified in the context of population-based genomic screening.

## Article information

### Ethics declaration

The Helix Research Network protocol has been IRB-approved, enabling secondary research use of data. Approvals for the protocol were granted by the Salus IRB (reliance on Salus for all sites; approval number 21143), the WCG IRB (Western Institutional Review Board, WIRB-Copernicus Group; approval number 20224919), the MUSC Institutional Review Board for Human Research (approval number Pro00129083), and the University of Nevada, Reno Institutional Review Board (approval number 7701703417). All participants provided written informed consent prior to participation. All data used for research had direct identifiers removed to safeguard participant privacy.

### Data sharing statement

Access to Helix Research Network^TM^ (HRN) data is available to qualified researchers subject to approval by the HRN Steering Committee and Helix. Interested researchers must enter into a Data Use Agreement, which prohibits re-identification of participants, sharing of data with third parties, and uploading data to public domains. The HRN is open to individual collaborations with scientific researchers. Considerations for data access requests include: (1) affiliation with an accredited academic institution that is committed to participant privacy and data security; (2) specificity, type and volume of data requested; (3) feasibility of the proposed research project; and (4) resource commitments from Helix and HRN member institutions required to support a collaboration.

## Data Availability

Access to Helix Research NetworkTM (HRN) data is available to qualified researchers subject to approval by the HRN Steering Committee and Helix. Interested researchers must enter into a Data Use Agreement, which prohibits re-identification of participants, sharing of data with third parties, and uploading data to public domains. The HRN is open to individual collaborations with scientific researchers. Considerations for data access requests include: (1) affiliation with an accredited academic institution that is committed to participant privacy and data security; (2) specificity, type and volume of data requested; (3) feasibility of the proposed research project; and (4) resource commitments from Helix and HRN member institutions required to support a collaboration.

## Acknowledgements

We thank all the participants of Ohio State Genomic Health, GeneConnect, Imagine You, DNA Answers, the Genetic Insights Project, the Healthy Nevada Project, In Our DNA SC, myGenetics, and The Gene Health Project. Funding was provided to the Healthy Nevada Project by the Renown Institute for Health Innovation and the Renown Health Foundation. The Healthy Nevada Project receives funding from Gilead Sciences, outside the scope of this research. Funding was provided to the myGenetics program by HealthPartners. Funding was provided to C.A.A.C. by the European Union’s Horizon Europe, under grant agreement No 101136962. Funding was provided to C.A.A.C. by UK Research and Innovation (UKRI) under the UK government’s Horizon Europe funding guarantee [grant agreements No 10098097, No 10104323]. Funding was provided to C.A.A.C. by Federal Department of Economic Affairs, Education and Research EAER, State Secretariat for Education, Research and Innovation SERI. We also acknowledge the entire Helix bioinformatics and lab teams for their contributions to the production of the exome sequencing pipeline as well as the research administration team for coordinating the project. We are grateful to the InSiGHT Hereditary Colorectal Cancer/Polyposis Expert Panel (InSiGHT VCEP) for providing such detailed and clear recommendations to interpret genetic variants in Lynch syndrome genes. Lastly, we thank Dr. Xiao-Fei Kong, Dr. Kevin Hughes, Dr. Raymond Kim, and Dr. Alan Yahanda for their valuable feedback and discussions related to this manuscript.

## References

1. Hampel H, Hall MJ: Hereditary Aspects of Colorectal Cancer: Mismatch Repair Genes Drive Lynch Syndrome [Internet]. Journal of the Advanced Practitioner in Oncology 9:311, 2018[cited 2024 Dec 18] Available from: https://pmc.ncbi.nlm.nih.gov/articles/PMC6333561/

2. Grzymski JJ, Elhanan G, Morales Rosado JA, et al: Population genetic screening efficiently identifies carriers of autosomal dominant diseases [Internet]. Nat Med 26:1235–1239, 2020Available from: 10.1038/s41591-020-0982-5

3. Buchanan AH, Lester Kirchner H, Schwartz MLB, et al: Clinical outcomes of a genomic screening program for actionable genetic conditions [Internet]. Genetics in Medicine 22:1874– 1882, 2020[cited 2025 Apr 22] Available from: https://www.nature.com/articles/s41436-020-0876-4

4. National Comprehensive Cancer Network: NCCN Guidelines Version 3.2024 Lynch Syndrome [Internet], 2024[cited 2025 Jan 13] Available from: https://www.nccn.org/home

5. Syngal S, Brand RE, Church JM, et al: ACG clinical guideline: Genetic testing and management of hereditary gastrointestinal cancer syndromes [Internet]. Am J Gastroenterol 110:223–62; quiz 263, 2015Available from: https://pmc.ncbi.nlm.nih.gov/articles/PMC4695986/

6. US Preventive Services Task Force, Davidson KW, Barry MJ, et al: Screening for colorectal cancer: US Preventive Services Task Force recommendation statement: US preventive services task force recommendation statement [Internet]. JAMA 325:1965–1977, 2021[cited 2024 Dec 18] Available from: https://jamanetwork.com/journals/jama/articlepdf/2779985/jama_davidson_2021_us_210011_1629739999.39667.pdf

7. Møller P, Seppälä TT, Bernstein I, et al: Cancer risk and survival in carriers by gene and gender up to 75 years of age: a report from the Prospective Lynch Syndrome Database [Internet]. Gut 67:1306–1316, 2018Available from: 10.1136/gutjnl-2017-314057

8. Dominguez-Valentin M, Sampson JR, Seppälä TT, et al: Cancer risks by gene, age, and gender in 6350 carriers of pathogenic mismatch repair variants: findings from the Prospective Lynch Syndrome Database [Internet]. Genetics in Medicine 22:15–25, 2019[cited 2025 Apr 22] Available from: https://www.nature.com/articles/s41436-019-0596-9

9. Dominguez-Valentin M, Haupt S, Seppälä TT, et al: Mortality by age, gene and gender in carriers of pathogenic mismatch repair gene variants receiving surveillance for early cancer diagnosis and treatment: a report from the prospective Lynch syndrome database [Internet]. EClinicalMedicine 58:101909, 2023Available from: 10.1016/j.eclinm.2023.101909

10. SEER*Explorer [Internet][cited 2025 Apr 22] Available from: https://seer.cancer.gov/statistics-network/explorer/application.html

11. McLaren W, Gil L, Hunt SE, et al: The ensembl variant effect predictor [Internet]. Genome Biol 17, 2016Available from: 10.1186/s13059-016-0974-4

12. Chen S, Francioli LC, Goodrich JK, et al: A genomic mutational constraint map using variation in 76,156 human genomes [Internet]. Nature 625:92–100, 2024Available from: 10.1038/s41586-023-06045-0

13. Ioannidis NM, Rothstein JH, Pejaver V, et al: REVEL: An ensemble method for predicting the pathogenicity of rare missense variants [Internet]. Am J Hum Genet 99:877–885, 2016Available from: 10.1016/j.ajhg.2016.08.016

14. Jaganathan K, Kyriazopoulou Panagiotopoulou S, McRae JF, et al: Predicting splicing from primary sequence with deep learning [Internet]. Cell 176:535–548.e24, 2019Available from: 10.1016/j.cell.2018.12.015

15. Jia X, Burugula BB, Chen V, et al: Massively parallel functional testing of MSH2 missense variants conferring Lynch syndrome risk [Internet]. Am J Hum Genet 108:163–175, 2021Available from: 10.1016/j.ajhg.2020.12.003

16. Shuey MM, Stead WW, Aka I, et al: Next-generation phenotyping: introducing phecodeX for enhanced discovery research in medical phenomics [Internet]. Bioinformatics 39:btad655, 2023Available from: https://academic.oup.com/bioinformatics/article/39/11/btad655/7335839

17. Davidson-Pilon C: lifelines: survival analysis in Python [Internet]. J Open Source Softw 4:1317, 2019Available from: 10.21105/joss.01317

18. Fummey E, Navarro P, Plazzer J-P, et al: Estimating cancer risk in carriers of Lynch syndrome variants in UK Biobank [Internet]. J Med Genet 61:861–869, 2024Available from: https://jmg.bmj.com/content/jmedgenet/61/9/861.full.pdf

19. Park J, Karnati H, Rustgi SD, et al: Impact of population screening for Lynch syndrome insights from the All of Us data [Internet]. Nat Commun 16:1–7, 2025Available from: https://www.nature.com/articles/s41467-024-52562-5

